# Are digital technology interventions effective to reduce loneliness in older adults? A systematic review and meta-analysis

**DOI:** 10.1101/2020.08.27.20183012

**Authors:** Syed Ghulam Sarwar Shah, David Nogueras, Hugo Cornelis van Woerden, Vasiliki Kiparoglou

**Affiliations:** NIHR Oxford Biomedical Research Centre, Oxford University Hospitals NHS Foundation Trust, John Radcliffe Hospital, Headington, Oxford, OX3 9DU, England, UK; Radcliffe Department of Medicine, University of Oxford, Level 6, West Wing, John Radcliffe Hospital, Headington, Oxford OX3 9DU, England, UK; EvZein Limited, Holley Crescent, Headington, Oxford, OX38AW, England, UK; Public Health Agency, 12 - 22 Linenhall Street, Belfast, BT2 8BS, Northern Ireland, UK; Centre for Health and Science, University of the Highlands and Islands, Old Perth Road, Inverness, IV2 3JH, Scotland, UK; Nuffield Department of Primary Care Health Sciences, University of Oxford, Radcliffe Observatory Quarter, Woodstock Road, Oxford. OX2 6GG, England, UK

## Abstract

**Objective:** To review the latest literature on the effectiveness of DTIs in reducing loneliness in (older) adults.

**Data Sources:** Electronic searches in PubMed, Medline, CINAHL, EMBASE and Web of Science covering publication period from 1 January 2010 to 31 July 2019.

**Subjects:** Adult men and women

**Design:** Systematic review and meta-analysis

**Main Outcome Measure:** Loneliness.

**Study Selection:** Primary studies that used DTIs for tackling loneliness in adults (aged ≥18 years) with follow-up measurements at least three months or more and publication in the English language.

**Data Extraction and Synthesis:** Two researchers independently screened articles and extracted data on several variables: participants, interventions, comparators and outcomes. Data was extracted on the primary outcome i.e. loneliness measured at the baseline and follow-up measurements at three, four, six and twelve months after the intervention.

**Results:** Six studies were selected from 4939 articles screened. Selected studies included 5 clinical trials (4 RCTs and 1 quasi experimental study) and one before and after study, which enrolled 646 participants (men =154 (24%), women =427 (66%), no gender information =65 (10%) with average age between 73 and 78 years (SD 6-11). Five clinical trials were included in the meta-analysis and standardised mean differences (SMD) were calculated for each trial and pooled across studies using a random effects model. The overall effect estimates were not statistically significant in follow-up measurements at three months (SMD= 0.02, 95% CI= −0.36, 0.40; P=0.92), four months (SMDs= −1.11, 95% CI= - 2.60, 0.38; P=0.14) and six months (SMD= −0.11, 95% CI= −0.54, 0.32; P=0.61). The quality of evidence was very low to moderate in these trials.

**Conclusions:** There is insufficient evidence to make conclusions that DTIs are effective in reducing loneliness in older adults. Future research may consider RCTs with larger sample sizes and longer duration of interventions and follow-up.

## 1 INTRODUCTION

Loneliness is an important public health problem (1), which has been seriously exacerbated because of recent isolation, social distancing, and lockdown measures for tackling the COVID-19 pandemic (2, 3). Thus, the burden of loneliness is expected to rise due to the COVID-19 crisis in countries (4) beyond developed countries where it is already high (5-12). The use of digital technology intervention have been suggested for mitigating the impacts of loneliness and attempting to deal with isolation due to social distancing in the epidemic situations such as the COVID-19 pandemic (3).

Loneliness refers to subjective feelings of an individual because of a perceived discrepancy between actual and the desired social relationships (13, 14). While loneliness affects people of all ages,(14, 15) older and vulnerable people are affected more (10, 16, 17). It is associated with the social and physical environment (16-18). Loneliness enhances the risk of poor mental and physical health (13, 19-22), dementia (23), premature mortality and all-cause mortality (20) particularly in older adults (22).

Loneliness could be addressed through a range of social (24) and technological interventions (25) such as digital applications (apps), online social networks, and social robots (26).

However, there is limited evidence on the effectiveness of technological interventions for loneliness (27). While several previously published reviews report that these interventions are effective in reducing loneliness (28-32), some of these studies are weak and have a high risk of bias (33). Some use very selected technological interventions and cover literature published over a very short time period i.e. between January 2010 and January 2013 (29). These findings suggest the need for further research (30, 31) to assess and identify the latest digital technological interventions that are effective in reducing loneliness (27, 32).

We therefore appraise the latest evidence to assess the effectiveness of digital technology interventions to reduce loneliness, which is imperative not only from the perspectives of patients and their families but also from the perspectives of other stakeholders such as public health, health and social care providers, and health insurers (34).

### 1.1 Study objectives

Our main objective was to assess the effectiveness of digital technology interventions (DTIs) to reduce loneliness in adults. The secondary objective was to identify DTIs that are used to reduce loneliness in adults.

### 1.2 Review questions

Our primary question was: “Are DTIs effective for reducing loneliness in adults?” and the secondary question was “What DTIs are used for reducing loneliness in adults?”

### 1.3 Outcome measures

Our main outcome measure was loneliness. We extracted loneliness scores measured at the baseline (before the intervention) and the follow-up measurements (at the end of intervention or sometime after the intervention) for the intervention group, and the control group(s) if any.

## 2 METHODS

### 2.1 Protocol registration and publication

We registered our systematic review and meta-analysis with the PROSPERO database on 10^th^ June 2019 (Registration ID CRD42019131524) (35) and we published our protocol (34).

### 2.2 Definition: Digital technology intervention

We defined a digital technology intervention as an intervention that applies digital technology i.e. the technology, equipment and applications that process information in the form of numeric codes, usually a binary code (36).

### 2.3 Eligibility Criteria

We selected studies that met our predefined eligibility Criteria (34). These included interventional studies (randomised and non-randomised) on loneliness using DTIs. The DTIs included the use of computers, computer tablets, iPads, Internet, online videos, communication, chatting, social groups, meetings, conferences and messages, sensors, (social) robots, (smart) phones, social media tools and the World Wide Web. A minimum Intervention duration and follow-up period was set at three months. Participants included both male and female adults (aged 18 years and more) living in different settings (i.e. residential dwellings including private residences and care / nursing homes in any country. The studies were limited to journal articles in the English language published from 1 January 2010 to 31 July 2019.

### 2.4 Information sources and keywords

We did electronic searches in PubMed, Medline, EMBASE, CINAHL and Web of Science covering the publication period from 1 January 2010 to 31 July 2019. We used an a priori list of keywords prepared in our preliminary literature searches (34). The keywords were of two categories: medical condition / problem (i.e. loneliness, lonely, isolation, aloneness, disconnect*, solitude, singleness*, lonesomeness, solitariness, and remoteness).and intervention / technology (i.e. digital, technolog*, sensor*, robot*, internet, social media, *phone*, online, iPad*, tablet*, computer*, electronic, web, video, and videoconference), as reported in our protocol (34).

### 2.5 Literature searches

First, we searched the keywords in the ‘subject headings’ such as Medical Subject Headings (MeSH) major terms in the PubMed or equivalent in other databases (Appendix-1). Thereafter, we searched for keywords in the ‘title’ and ‘abstract’ fields in the selected databases using three Boolean operators: ‘OR’, ‘AND’ and ‘NOT’. In addition, we hand searched through reference lists of shortlisted articles. We wrote emails to the authors of two studies for full copies of their research articles (37, 38), which were gratefully provided to us. We contacted the authors of two further studies for missing / additional data (39, 40). We had good response from the authors of both studies and data were thankfully provided for one study (40). We sought support from an expert librarian for running literature searches.

### 2.6 Study Selection

Searches retrieved 4939 articles that included 965 duplicate articles, which were removed (Fig. 1). Two researchers (SGSS and DN) independently screened titles of 3974 articles and read abstracts of 442 articles, which led to exclusion of 3876 articles.. Full text of the remaining 98 articles was read independently by three researchers (SGSS, DN and VK). When recommendations differed between reviewers at the title, abstract and full text review stages, another reviewer (HCvW) reviewed these articles and his recommendations to either include or exclude an article were final. Finally, we excluded 92 articles and included six articles in the data-extraction and narrative synthesis, and included five studies in the meta-analysis (Fig. 1).

**Fig. 1.**
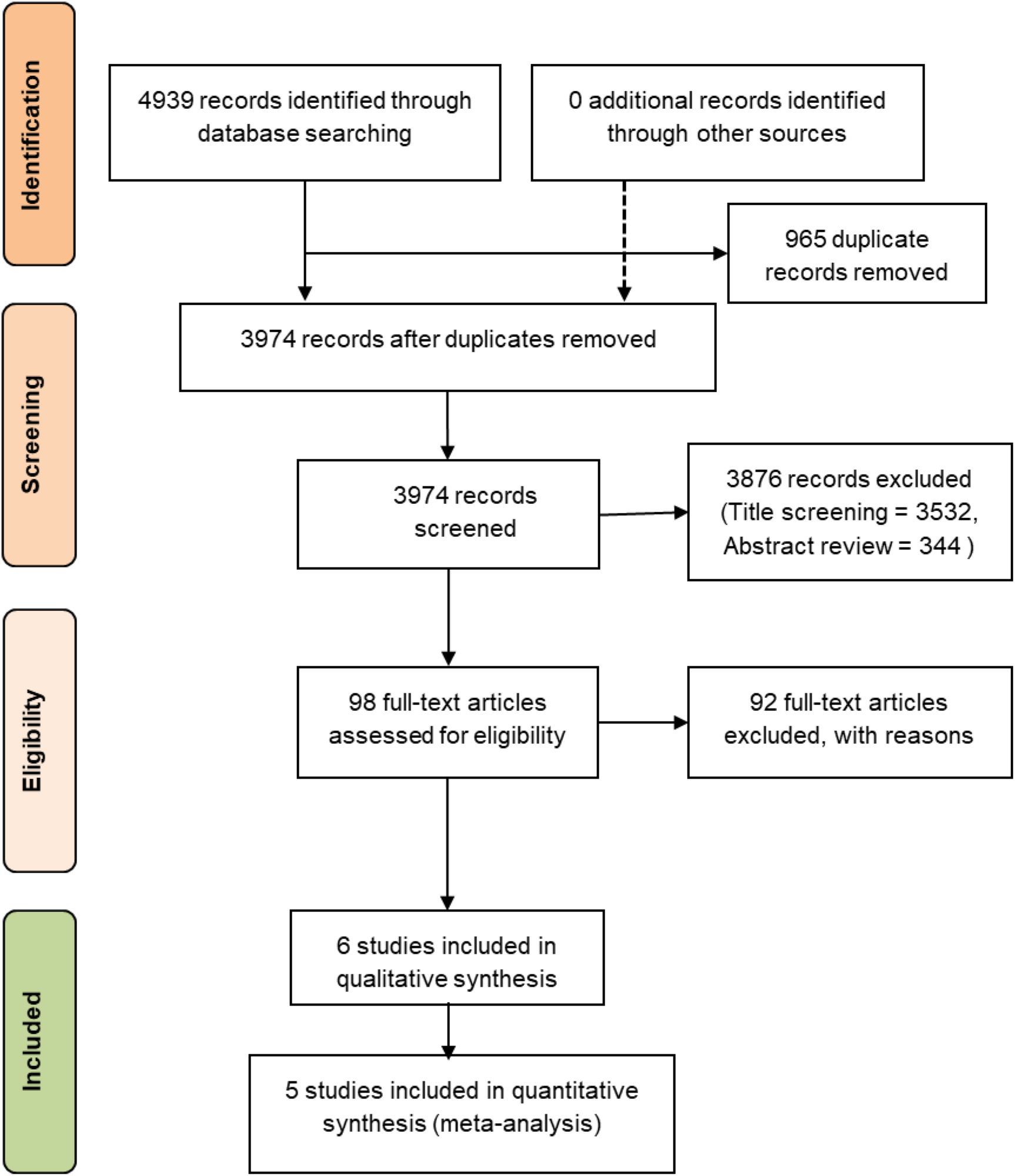
PRISMA Study selection flow diagram

### 2.7 Data collection process

For data collection, we used a priori data extraction template (Tables 1-2), which comprised a number of columns: Author(s), year and country of study; study aim/objectives; research design; settings; participants’ characteristics (age, gender and ethnicity); health/medical condition; sampling method and size; participant attrition (numbers / %), research method(s) / data collection tool(s); intervention(s) (e.g. type/tool of digital technology), comparator(s).(e.g. alternative intervention, placebo or care as usual), intervention duration (weeks / months),.measurement stages (e.g. baseline and follow-up - weeks/months after the baseline), outcomes/results/findings (e.g. loneliness scores (including statistics e.g. mean values, standard deviations, standard errors and confidence intervals) and authors’ conclusion(s) (34).

**Table 1.** Characteristics of included studies

| Authors<br>(publication<br>year) country | Quality of<br>Evidence | Research<br>Design | Settings | Participants |  |  | Main Health<br>/ Medical<br>condition(s) | Sampling<br>method | Sample size |  |  | Participant<br>Attrition | Research<br>Method(s)/<br>Data<br>collection<br>tool(s) |
| --- | --- | --- | --- | --- | --- | --- | --- | --- | --- | --- | --- | --- | --- |
|  |  |  |  | Age (years) | Gender | Ethnicity |  |  | Total | Intervention<br>group | Control<br>group /<br>end of<br>study |  | Scale used<br>for<br>measuring<br>loneliness |
| Tsai et al<br>(2010) (64)<br>Taiwan | Medium | Quasi-<br>experimental<br>study (N-RCT) | Nursing<br>home | Baseline:<br>Experimental<br>group:<br>average age =<br>74.2 (SD<br>10.18);<br>Control group:<br>average age =<br>78.48 (SD<br>6.75) | Male = 24<br>(experimental<br>group =10,<br>control group<br>=14);<br>Female = 33<br>(experimental<br>group =14,<br>control group<br>=19) | Not reported<br>(probably all<br>Taiwanese /<br>Chinese) | Loneliness<br>and<br>Depression | Purposive | 57 =<br>Baseline,<br>49 = End<br>of study | 24 =<br>Baseline,<br>21 = Follow-up | 33 =<br>Baseline<br>, 28 =<br>Follow-up | Total = 8 (5<br>from control<br>group and 3<br>from<br>experimental<br>group),<br>attrition rate<br>= 14% | UCLA<br>Loneliness<br>Scale<br>(Russel et<br>al, 1980) |
| van der Heide<br>et al (2012)<br>(56) The<br>Netherlands | Low | Before and<br>after study<br>(with<br>intervention<br>group only,<br>no control<br>group) | Elderly<br>home care | Baseline:<br>average 73.2<br>(SD 11.8),<br>range 32-90;<br>End of study<br>stage:<br>Average 73.1<br>(SD 11.2),<br>range 38-90 | Baseline: Male =<br>26 (30.2%),<br>Female =60<br>(69.8%), Missing<br>values n=44;<br>End of study:<br>Male = 25<br>(29.4%), Female<br>= 60 (70.6%),<br>missing<br>values.=0 | Not reported | Feeling of<br>loneliness<br>and safety | Convenienc<br>e | 130 | 130 | intervent<br>ion<br>group at<br>the end<br>of study<br>= 85.<br>No<br>control<br>group | Total =45,<br>attrition rate<br>=34.6% | De Jong-Gierveld<br>loneliness<br>scale (Score<br>range: 0-11) |
| Larsson et al<br>(2016) (44)<br>Sweden | High | Randomise,<br>crossover<br>study | Living in<br>ordinary<br>housing<br>without any<br>home care<br>services | Total sample:<br>range: 61-89<br>(SD 71.2);<br>Group 1<br>(intervention /<br>control group):<br>range 66-89<br>(SD 73.6);<br>Group 2<br>(Control/interv<br>ention group):<br>range 61-76<br>(SD 69.0) | Total: male =6,<br>female = 24, (3<br>males and 12<br>females each in<br>Group 1<br>(intervention /<br>control group)<br>and Group 2<br>(Control /<br>intervention<br>group) | Not reported<br>(probably all<br>Swedes) | Loneliness | Randomised<br>(after<br>recruitment) | 30 | 15 =<br>Baseline,<br>14 = Follow-up | 15 =<br>Baseline<br>, 14 =<br>Follow-up | Total = 2 (1<br>participant<br>each from<br>intervention<br>and control<br>groups),<br>attrition rate<br>= 6.7% | UCLA<br>loneliness<br>scale<br>(Russell,<br>1996) - the<br>Swedish<br>version<br>(Engelberg<br>and Sjoberg<br>(2005)<br>(20 items,<br>score range<br>20-80) |
| Czaja et al (2018) (40)<br>United States | High | Multisite randomised controlled trial | Living in independent housing in the community | Baseline: total sample mean = 76.15 (SD 7.4), range: 65-98; Intervention (PRISM) group: mean = 76.9 (SD 7.3); Control (Binder) group: mean = 75.3 (SD 7.4) | Baseline: Female = 78% (number not reported), Male = 22% (number not reported); PRISM/Intervention group: Female 79.3% (n=119); Binder (control) group: Female 76.7% (n=115) | Baseline: White 54% and Non-white (46%); PRISM/Intervention group: Non-white/Hispanic n=12, 8%; Binder group: Non-white/Hispanic n=15, 10.0% | Social isolation, Social support, Loneliness, and Wellbeing | Randomised | 300 (150 in each intervention arm (PRISM group) and control arm (Binder group)) | 150 = Baseline, 134 = Follow-up | 150 = Baseline, 118 = Follow-up | Total = 56, (45 at 6 months and 11 at 12 month follow-up), Attrition rate = 18.7% | UCLA Loneliness Scale (Russell, 1996) (score range 20-80) |
| Morton et al (2018) (63)<br>United Kingdom | High | 2 (condition: training, control) × 2 (population: domiciliary, residential) × 2 (time: baseline, follow-up) design | Receiving care in own home / supported housing in the community ("domiciliary care"), or residential care in care homes | Female: mean = 80.71 (SD =8.77) Male: data not reported | Follow-up: Total = 76; Female = 50, Male = 26 | Not reported | Wellbeing and social support | Randomised | 97 = Baseline, 76 = Follow-up | 53 = Baseline, 44 = Follow-up | 44 = Baseline, 32 = Follow-up | Total = 21 (9 = experimental group, 12 = control group) attrition rate = 21.6% | UCLA Loneliness Scale (Russell, 1996) (score range 20-80) |
| Jarvis et al (2019) (62)<br>South Africa | High | Randomised control study | Inner-city residential NGO care facilities for resource-restricted older people (≥60 years) | Mean = 74.93 (SD 6.41), range = 61-87 | Baseline: Male = 6 (18.8%); Female = 26 (81.2%) | Mostly Asian / Indian, numbers not reported | maladaptive cognitions and loneliness | Randomised | 32 = Baseline (intervention group =15, control group =17), Final = 29 (intervention group =13, control group =16) | 15 = Baseline, 13 = Follow-up | 17 = Baseline, 16 = Follow-up | Total = 3 (2 = intervention group, 1 = control group), Attrition rate = 15.6% | De Jong-Gierveld loneliness scale (Score range: 0-11) |

**Table 2.** Interventions and outcome measurements in included studies

| Study | Intervention(s) | Comparator(s) | Intervention duration | Follow-up duration | Outcomes: Loneliness scores by measurement stages |  |  |  |  | Results / Findings | Authors' Conclusion |
| --- | --- | --- | --- | --- | --- | --- | --- | --- | --- | --- | --- |
|  |  |  |  |  | Mean Scores (SD) |  |  |  |  |  |  |
|  |  |  |  |  | Baseline | 3 months | 4 months | 6 months | 12 months |  |  |
| Tsai et al (2010)(64) | Videoconference interaction (using either MSN or Skype) | Regular care | 3 months | 3 months | Intervention group = 50.58 (SD 11.16), Control group = 46.55 (SD 9.07) | Intervention group = 47.33 (SD = 13.50); Control group = 46.68 (SD = 9.08) | Not measured | Not measured | Not measured | Loneliness: Intervention group mean (SD) = baseline 50.58 (11.16), one week 49.75 (11.79) and 3 months 47.33 (13.50); Control group mean (SD) = baseline 46.55 (9.07), one week 47.06 (8.75) and 3 months 46.68 (9.08); Differences between groups were compared at three points (baseline, one week, and three months) using multiple linear regression of the generalized estimating equations. Unadjusted / fixed effect of effectiveness of videoconferencing intervention videoconference vs. control: at one week Beta ($\beta$) = -1.21. SE 0.50, $\chi^2$ =5.95, p = 0.02 and at 3 months Beta ($\beta$) = -2.84, SE 1.28, $\chi^2$ = 4.89, p = 0.03. | Videoconferen<br>ce program<br>alleviate<br>depressive<br>symptoms and<br>loneliness in<br>elderly<br>residents in<br>nursing homes |
| van der Heide et al (2012)(56) | CareTV intervention including Caret duplex video/voice network | No control group and no comparator | 12 months | 12 months | Interventio<br>n group = 5.97 (SD 2.77); No control group | Not measured | Not measured | Not measured | Interventio<br>n group = 4.02 (SD 3.91); No control group | Group level total loneliness: Inclusion stage: mean 5.97 (SD 2.77) (scale 0-11), End of study: mean 4.02 (SD 3.91), p = 0.001. Individual level total Loneliness: total loneliness decreased in 54 out of 85 participants (equally lonely 11, more lonely 20 and less lonely 54 individual participants) | CareTV intervention decreased the feeling of loneliness in the participants; however, participants were feeling moderate loneliness at the end of the study |
| Larsson et al (2016)(44) | Social internet-based activities (SIBAs) such as Skype and Facebook | No comparator intervention reported | 3 months | 34 weeks (3 months exposure to each group) | Group 1 (I/C group) = 45.53 (SD=7.41); Group 2 (C/I group) = 43.93 (SD 8.61) | Group 1 (I/C group) = 42.43 (SD=7.44); Group 2 (C/I group) = 41.93 (SD 8.82), | Not measured | 3 months after cross over: Group 1 (I/C group, No intervention) = 42.0 (SD=7.34); Group 2 (C/I group, Intervention introduced) = 39.50 (SD 10.42). | Not measured | % change between T2 and T1: Group 1 = mean score : -0.07% (SD 0.07), p. 0.003; Group 2 = mean score : -0.05% (SD 0.09), p. 0.049; % change between T3 and T1: Group 1 = mean score : -0.08% (SD 0.08); Group 2 = mean score : -0.09% (SD 0.13); Comparison of pre and post-intervention scores group 1 = p. 0.003 and group 2 = p. 0.049 | SIBA interventions have potential to reduce experiences of loneliness in socially vulnerable older adults. |
| Czaja et al (2018)(40) | Personal Reminder Information and Social Management (PRISM) system | A notebook with printed content similar to that within PRISM group (intervention group) - include a Lenovo "Mini Desktop" PC with a keyboard, mouse (or trackball for those who were unable to control a mouse), a 19" LCD monitor, the PRISM software application, and a printer and internet access. | 12 months | 12 months | Intervention (PRISM) group = 39.8 (SD 9.7); Control (Binder) group = 40.2 (SD 10.3) | Not measured | Not measured | Intervention (PRISM) group = 37.8 (SD 9.54); Control (Binder) group = 40 (SD 10.62) | Intervention (PRISM) group = 36.9 (SD 9.16); Control (Binder) group = 38.43 (SD 9.37) | Baseline: Loneliness = PRISM group mean score 39.8 (SD 9.7) and Binder group mean score 40.2 (SD 10.3), follow-up at 6 months PRISM group about 37.8 and Binder group 39.6; Follow-up at 12 months PRISM group 36.9 and Binder group 38.3. | Technology applications such as PRISM may enhance social connectivity and reduce loneliness among older adults. |
| Morton et al (2018)(63) | “EasyPC” - a customized computer platform with a simplified touch-screen interface (Intervention group) | Care as usual (plus regular carer visits) | 3 months | 3 months | Intervention (Training) group (total of residential and domiciliary groups): =1.95 (SD: 0.73); Control group (total of residential and domiciliary groups): =2.08 (SD 0.80) | Not measured | Intervention (Training) group (total of residential and domiciliary groups): 1.86 (SD 0.66); Control group (total of residential and domiciliary groups): 2.12 (SD 0.62) | Not measured | Not measured | Loneliness scores= mean (standard error SE): Control group:- Residential group T1 = - 2.13 (SE .18), T2 =2.20 (SE .17), Domiciliary group T1 = 2.02 (SE .16), T2 = 2.05 (SE .15), total T1 = 2.08 (SE .12) and T2 = 2.12 (SE .11). Intervention (Training) group: Residential group: T1= - 1.95 (SE .16), T2 = 1.92 (SE .16), Domiciliary group: T1 = 1.89 (SE .13), T2 = 1.79 (SE .13), Total T1 = 1.92 (SE.10), T2 = 1.86 (SE .10). | Internet access and training can support the self and social connectedness of vulnerable older adults and contribute positively to well-being. |
| Jarvis et al (2019)(62) | Living In Network Connected Communities (mLINCC) WhatsApp group for low-intensity Cognitive Behavior Therapy (LI-CBT) intervention | Usual care, a separate WhatsApp group (mLINCC 2) | 3 months | 3-4 months | Not measured | Intervention group = 2.31 (SD = 1.33); Control group = 2.47 (SD = 2.1) | Intervention group = 1.38 (SD = 1.33); Control group = 4.0 (SD = 1.32) | Not measured | Not measured | loneliness levels: Total = T0–T1–T2, $\chi^2 = 14.62$, $p = 0.001$) | LI-CBT mHealth supported by the social networking platform of WhatsApp (mLINCC) showed significant improvements in loneliness and maladaptive cognitions. |

SGSS and DN independently extracted data from all included studies (n=6) using the data extraction template (Tables 1-2) and resolved discrepancies in extracted data with discussion. We compared and contrasted data extraction forms; thus, we avoided bias and reduced errors in the data extraction process (41). We extracted aggregated data at the study level as much as possible with respect to the intervention, which is imperative for the reproducibility of effective interventions (42, 43). Following suggestions for reporting data once from studies with duplicate and multiple publications,(41) we extracted, and report, data only once (44) from a research study with multiple publications (44, 45).

### 2.8 Data Synthesis and reporting

We report both a narrative synthesis (narrative summary) and a statistical (quantitative) synthesis (meta-analysis) of our review study as suggested for reporting of a systematic review on the effectiveness (46). In the narrative synthesis, we include all six studies and report their characteristics including the study design, settings, sample sizes, data collection methods, participants, intervention(s), comparator(s), outcome measurements and study conclusion(s)..

In the meta-analysis, we included five studies and pooled extracted data on loneliness measured by continuous loneliness scales.

In meta-analysis, the standardised mean difference (SMD) as a summary statistic for reporting continuous data has been suggested for studies that assess the same outcome but use different scales for measuring the outcome (47). In RevMan, the SMD is the effect size known as Hedges’ (adjusted) g, which is akin to Cohen’s d and it includes an adjustment for small sample size bias (47). In our review, the main outcome i.e. loneliness, was measured using different loneliness scales which included the UCLA loneliness scale (score range: 20-80) (48, 49) and the de Jong Gierveld loneliness scale (score range.0-11) (50).

We calculated the standardised mean values (SMDs) from extracted data i.e. loneliness mean scores with standard deviations (SD) and sample sizes in the intervention and control groups during the follow-up measurements at three, four and six months after the intervention. We conducted meta-analysis in the Review Manager (RevMan), version 5.3.5 software (51). In the meta-analysis, we ran the random effects model as the statistical model because we hypothesised that the true effect sizes between studies would vary (52, 53) due to differences in the methodological and clinical characteristics between studies (54) such as differences in the sample sizes, participants, interventions types and duration, and the follow-up measurement times.

The Cochrane guidelines (47) suggest that different study designs should not be combined in the meta-analysis because it can increase heterogeneity and studies with repeated measurements at different follow-up periods cannot be combined without a unit-of-analysis error (47). We therefore conducted meta-analyses for similar study designs i.e. clinical trials but separated them based on the follow-up measurement periods. Thus, we undertook a separate meta-analysis for each follow-up measurement at three, four and six months after the intervention. In addition, we ran meta-analyses when there were at least two or more studies for the same outcome or the same follow-up period (55). We therefore did not run a meta-analysis for follow-up measurements at 12 months reported in two studies because they involved different study designs i.e. an RCT with intervention and control groups (40) and a pre and post intervention study with only intervention group(56) to avoid increase in the heterogeneity (47). Thus, we also avoided the overestimation of the effect of intervention in the absence of a control group (57) in the pre and post intervention study (56). In addition, we did not conduct sensitivity analyses due to the small number of studies in meta-analyses at each follow-up point (55). We had only one pre-post study and we did not run meta-analysis for this study because meta-analysis cannot be run with only one study (47).

### 2.9 Assessment of research quality, bias and heterogeneity

We assessed the quality of research (Table 3) by applying the GRADE (Grading of Recommendations Assessment, Development and Evaluation) approach (58).

**Table 3.**
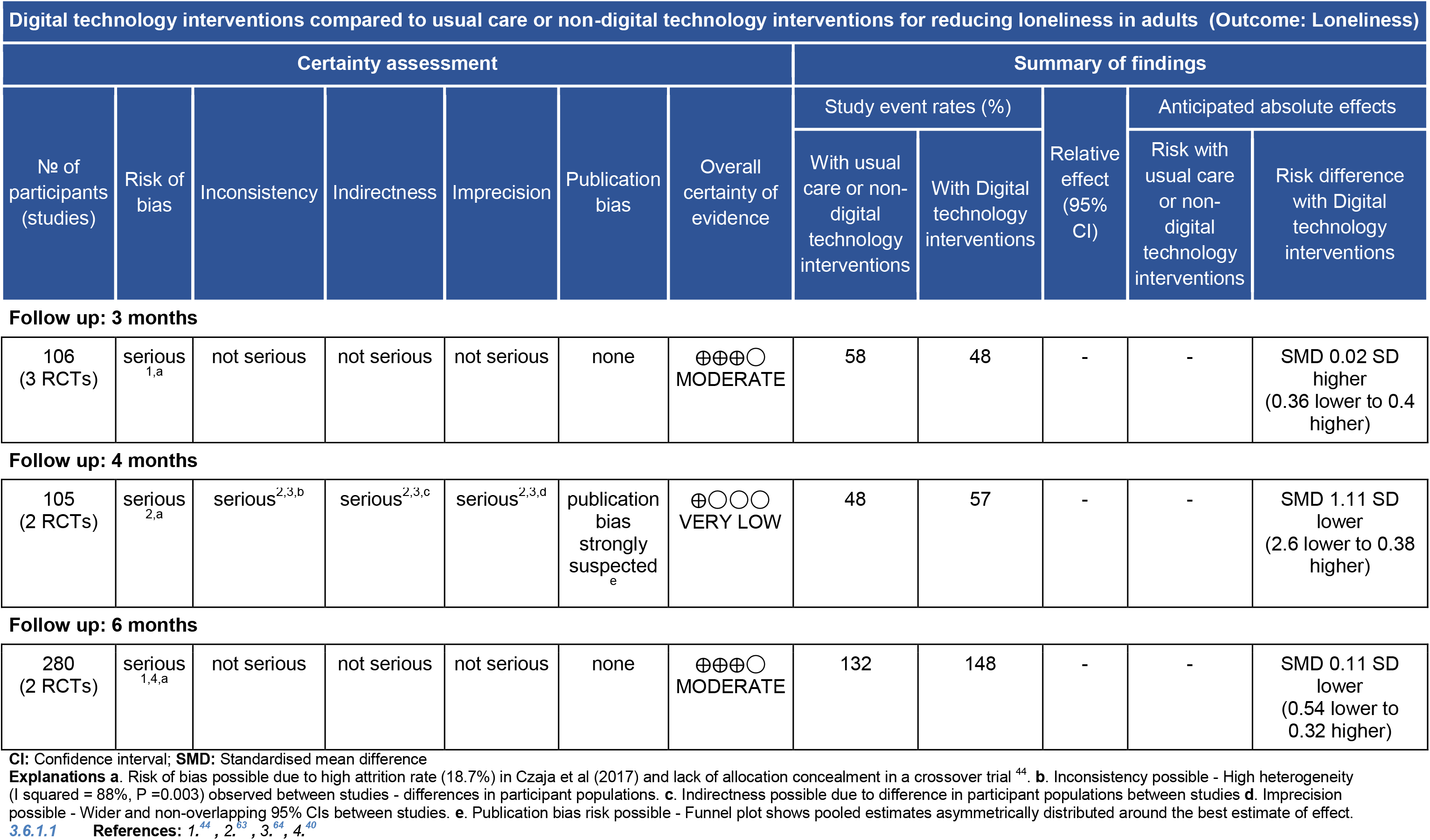
GRADE Quality of Evidence

We assessed risk of bias focusing on five domains: evaluation of sequence generation, allocation concealment, blinding (outcome assessors), incomplete data, selective outcome reporting, and assessing other biases using the Cochrane method (47). In a meta-analysis, publication bias can be assessed by graphical method using Funnel plots (47, 59) and statistical methods such as the Egger’s test (47); however, the both methods require at least 10 studies in the meta-analysis (47). When the number of studies is less, the Egger’s test has low power and it fails to differentiate chance from real asymmetry.(47). Similarly, assessing publication bias by Funnel plots with fewer studies would be of very limited usefulness because it would be difficult to spot the publication bias. We had three studies maximum in a meta-analysis (Fig. 2); hence, we could not check the publication bias by either method.

We checked heterogeneity i.e. variation in study outcomes / effect sizes between studies by the Cochran’s Q (χ^2^) Test with a significance level of *ρ* < 0.05 (25, 60). We used I^2^ statistics for determining the magnitude of heterogeneity (i.e. the proportion of variance in the true effect sizes) between studies (25). We considered I^2^ of ≤25%, between 25% and 50% and >50% as low (55), moderate (25, 61) and high heterogeneity between studies, respectively (19, 61).

### 2.10 Summary measures

We report the findings of our meta-analyses using the SMDs with 95% confidence intervals (CI) as a statistical summary with the forest plots (47).

## 3 RESULTS

### 3.1 Narrative synthesis

Participants, intervention(s), comparator(s), outcome measurements and study conclusion(s) are presented in Table 1. Characteristics of studies including the study designs, settings, sample sizes, data collection methods are given in Table 2.

#### 3.1.1 Study selection

The database search of PubMed, Medline, CINAHL, EMBASE, and Web of Science generated a total of 4939 papers (Fig. 1) of which six studies met the eligibility Criteria. All six studies are included in the narrative synthesis and five studies are included in the meta-analysis.

#### 3.1.2 Study Participants

The total number of participants enrolled in all six included studies was 646 (mean = 108, SD = 102, median = 77, IQR= 32 to 130). Studies varied in total sample sizes (mean 108, SD 102, range 30-300) and the sample sizes of intervention groups and control groups also varied at both the baseline and the follow-up measurements (Table 1). The attrition rate also varied between studies (range 7-35%, mean 19%, SD 10%).

Participants’ average age was between 73 and 78 years (SD 6-11). Total enrolled participants included 427 women (66%) and 154 men (24%) while no information about the gender was available for 65 (10%) participants. Studies varied by gender of participants i.e. female (mean= 66%, SD =16%, range = 46-81%) and male (mean =25%, SD =9%, range = 19-42%). Only two studies reported on participants’ ethnicity, which included white (54%) and non-whites (46%) in the US study (40) and mostly Asian Indians (no numbers reported) in the South African study (62).

#### 3.1.3 Study characteristics

The characteristics of included studies (n=6) are given in Tables 2. Of six studies included, four studies were randomised clinical trials (RCTs) (40, 44, 62, 63), one non-randomised clinical trial (NRCT) (64) and one pre- and post-test (before and after) study with intervention group only (no control group) (56) (Table 1).

#### 3.1.4 Study settings

Four studies were conducted in developed countries i.e. the Netherlands (56), UK (63), US Czaja et al., 2018) and Sweden (44). Two studies were undertaken in developing countries i.e. Taiwan (64) and South Africa (62).

The settings included living in independent housing in the community (40), living in ordinary housing without any home care services (44), receiving care in their own home/supported housing in the community (“domiciliary care”), or residential care in care homes (63), residential care facilities for older people (62), nursing home (64) and elderly home care (56).

Participants were selected by random sampling in four (60%) studies (40, 44, 62, 63) and the other two (20%) studies used purposive (64) and convenience (56) sampling each.

#### 3.1.5 Digital technology interventions

Digital technology interventions included social internet-based activities (SIBAs) such as Skype and Facebook (44), videoconferencing (64), customized computer platform with simplified touch-screen interface (63), personal reminder information and social management system (40), WhatsApp group (62) and video/voice network (56).

#### 3.1.6 Duration of intervention and measurement of main outcome measure

The duration of intervention was three months in four studies (44, 62-64) and 12 months in two studies (40, 56). The main outcome measure i.e. loneliness was measured at the baseline and multiple follow-up times, which included at three months in three studies (44, 62, 64), four months in two studies (62, 63), six months in two studies (40, 44) and 12 months in two studies (40, 56). Loneliness measurement tools were the UCLA loneliness scale (48, 49) used in four studies (40, 44, 63, 64) and the De Jong-Gierveld loneliness questionnaire (50, 65) used in two studies (56, 62). Table 2 presents loneliness scores measured in the intervention and control groups, if any, at the baseline and follow-ups. Narrative synthesis showed that there was reduction in loneliness in the intervention groups at the follow-ups compared to at the baseline (Table 2). A statistical summary of the loneliness measurements in the intervention and control groups at the follow-ups is reported in the meta-analysis section.

### 3.2 Meta-analysis

We ran three meta-analyses one each for follow-up measurements at three, four and six months involving three, two and two studies respectively.

#### 3.2.1 Meta-analysis for follow up measurements at 3 months

Three studies (44, 62, 64) involving 106 participants with follow-up measurement at three months were entered into a meta-analysis, which showed a very small reduction in loneliness in favour of the control (SMD = 0.02, 95%CI −0.36, 0.40) but it was not statistically significant (Z =0.10, P=0.92). The heterogeneity between studies was not statistically significant (τ^2^ =0.00, χ*^2^* = 0.10, P=0.95, I^2^ =0%) (Fig. 2 - panel A).

### 3.3 Meta-analysis for follow up at 4 months

Two studies (62, 63) involving 105 participants with four month follow-up were entered into a meta-analysis, which revealed a large reduction in loneliness in favour of the intervention (SMD = −1.11, 95%CI −2.60, 0.38) but was not statistically significant (Z=1.46; P=0.14).There was statistically significant high heterogeneity between studies (τ^2^ =1.03, χ^2^ = 8.84, P=0.003, I^2^ =88%) (Fig. 2 - panel B).

### 3.4 Meta-analysis for follow up measurements at 6 months

A meta-analysis involving 2 studies (40, 44) with 208 participants with six month follow-up showed a very small reduction in loneliness in favour of the intervention (SMD = −0.11, 95%CI −0.54, 0.32) but it was not statistically significant (Z=0.51, P=0.61). There was moderate heterogeneity between studies but not statistically significant (τ^2^ =0.05, χ^2^ = 1.58, P=0.21, I^2^ =37%) (Fig. 2- panel C).

**Fig. 2.**
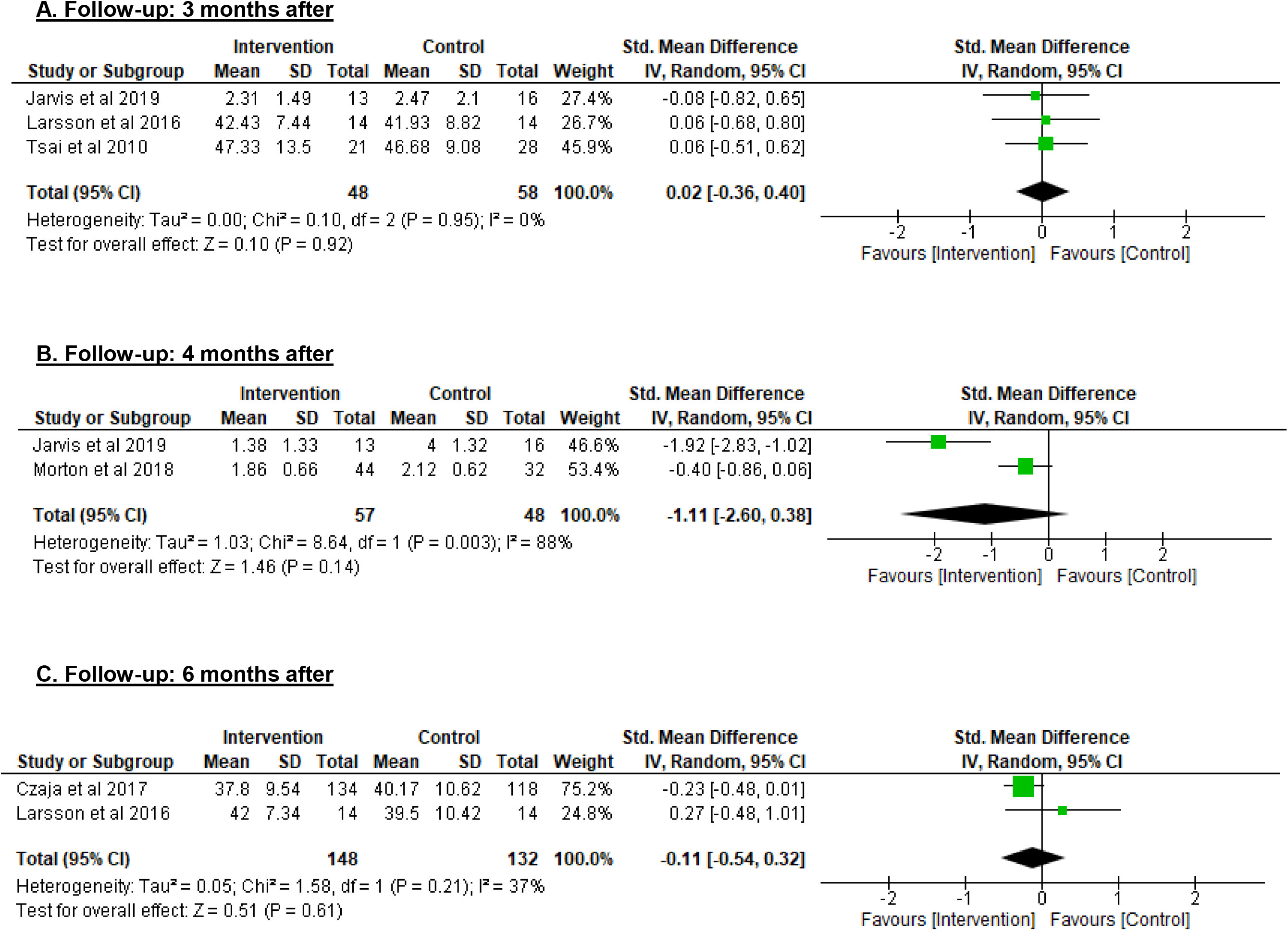
**Forest plots Comparison:** Digital technology Intervention vs Control, Outcome: Loneliness

### 3.5 Risk of Bias

Risk of bias assessment is presented in Figs. 3-4. A high risk of bias was noted more in the attrition bias and other bias; unclear risk of bias was detected in blinding of outcome assessment, allocation concealment and blinding of participants and personnel while a low risk of bias was observed especially in the random sequence generation and selective reporting (Fig. 3). In addition, most studies reported only within-group changes and not between-group comparisons of change, which may suggest a weak quality of the reporting of results and the analysis in these studies (Table 2).

**Fig. 3.**
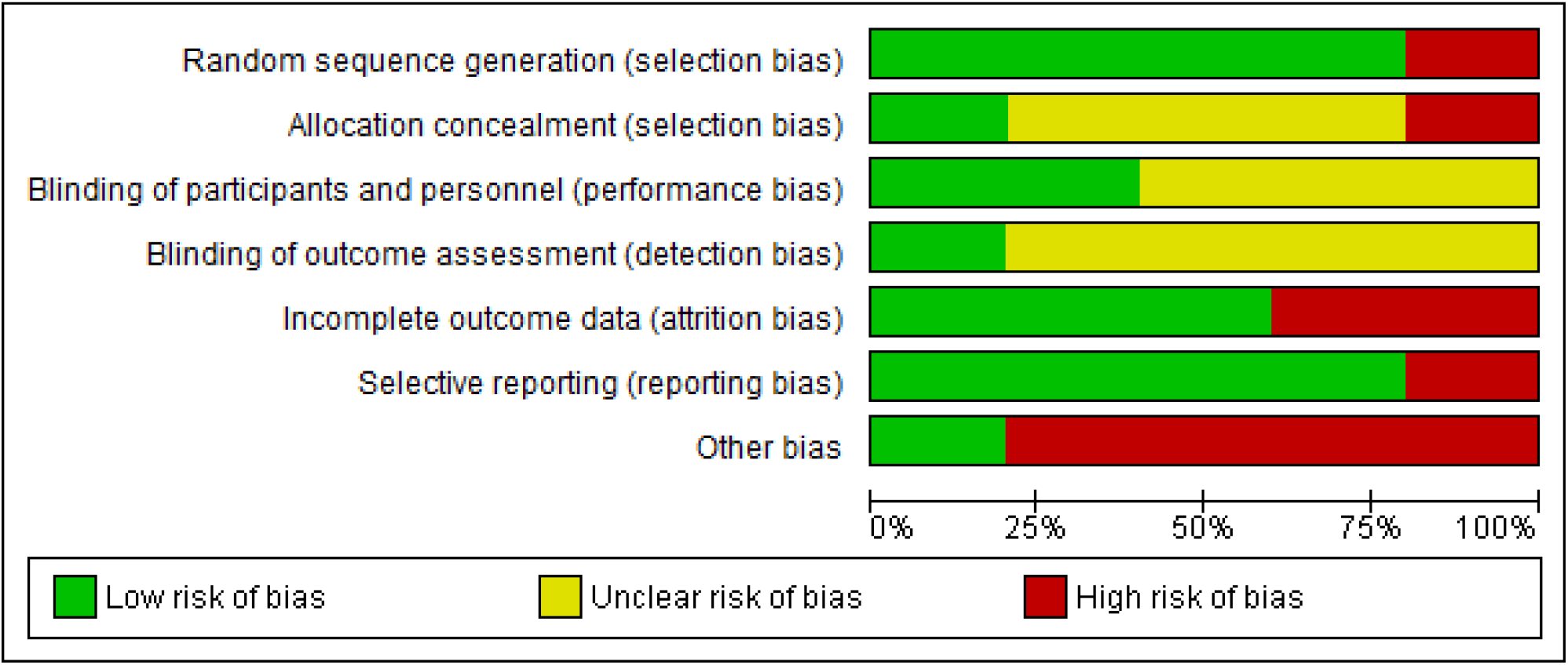
**Risk of bias graph:** Review authors’ judgements about each risk of bias item presented as percentages across all included studies

**Fig. 4.**
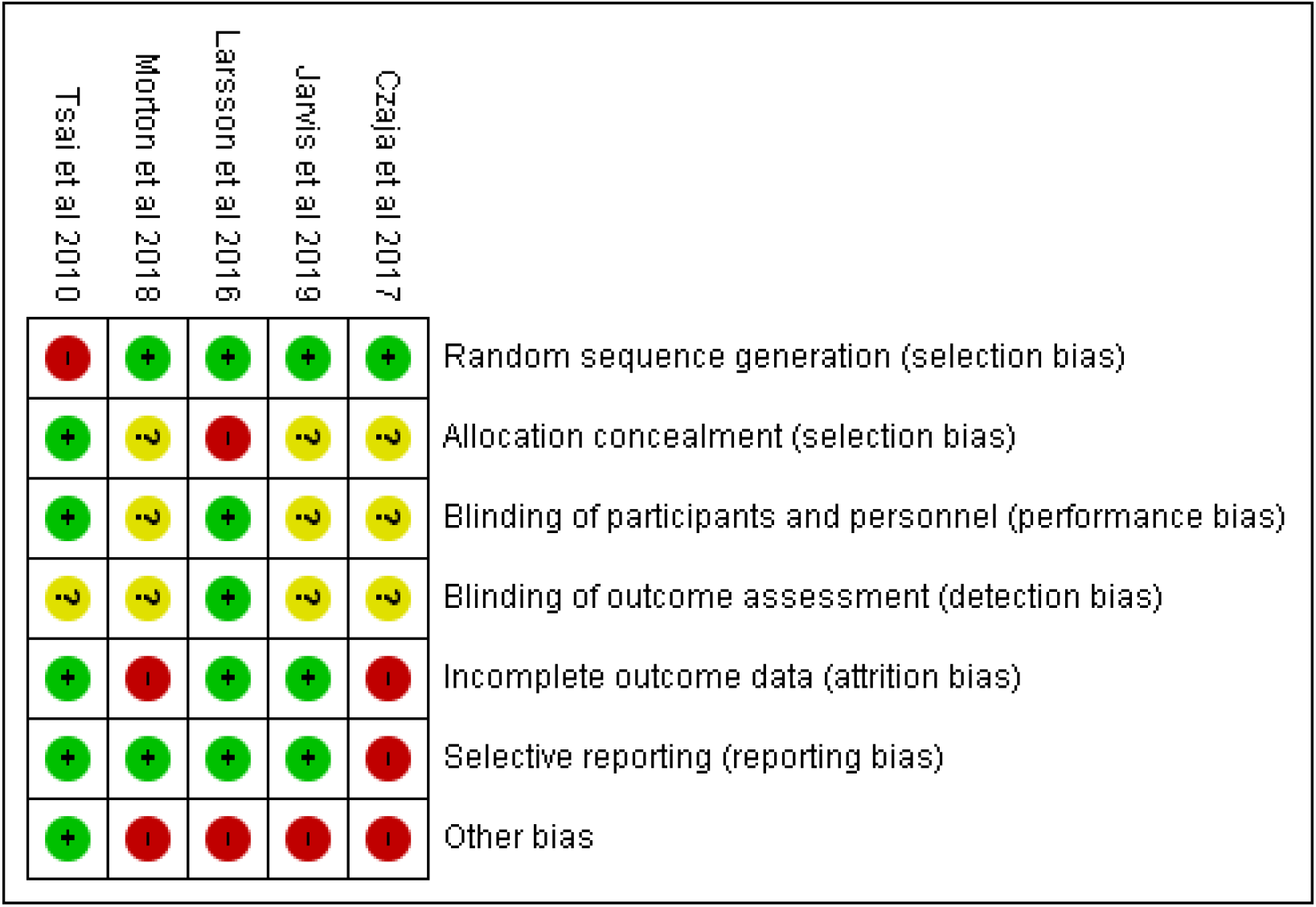
**Risk of bias summary:** Review authors’ judgements about each risk of bias item for each included study.

### 3.6 Quality of Evidence

We assessed the quality of evidence as moderate, very low and moderate in meta-analyses involving three (44, 62, 64), two (62, 63) and two studies(40, 44) with follow-up at three, four and six months respectively (Table 3 - GRADE quality of evidence).

## 4 DISCUSSION

To provide the answer whether digital technology is effective in reducing loneliness in adults, we appraised peer reviewed empirical research involving the application of DTIs in adults with loneliness. Our systematic review includes a narrative summary of six studies, which reported reduction in loneliness in the intervention groups at the follow-up compared to the baseline (Table 2). However, our meta-analyses of five clinical trials with follow-up measurements at three, four and six months showed no statistically significant pooled effect estimates as SMDs – the preferred method for summarizing effects on continuous outcomes such as loneliness. Although not statistically significant, the summary effect size at four months follow-up was better compared to the effect size at three and six months follow-up (Fig. 2).

Our meta-analysis also revealed that CIs of the summary effects of two studies i.e. (44). and (64) were very wide and the SMDs from these studies were more in favour of control group rather than the intervention group (Fig. 2). Thus, the wide widths of CIs of the summary effects in these studies leave uncertainty about the beneficial effect or otherwise of DTI on measures of loneliness.

Overall, findings of our meta-analysis provide insufficient evidence to make conclusions that DTIs are effective in reducing loneliness in (older) adults.

### 4.1 Summary of evidence

The quality of evidence of included studies was very low to moderate (Table 3) and there was a high heterogeneity between studies (62, 63). All included studies had a high proportion of female participants. Most notably, the total number of participants was low (44, 62) and the sample sizes were reduced further due to a high attrition rate in some studies (40, 63). The types and methods of DTIs varied between studies, which were conducted in diverse settings and different countries (Tables 1-2). These factors could have contributed in pooled estimates being not statistically significant.

There a few published meta-analyses on technological interventions for tackling loneliness covering literature published up to 2009 (25) and 2011 (28) whereas our review and meta-analysis includes the latest evidence published between 1January 2010 and 31July 2019. We did not replicate the findings of earlier meta-analyses that reported evidence suggesting technological interventions resulting in decreased loneliness (25, 28). A meta-analysis by (28) reported statistically significant evidence suggesting internet and computers reduce loneliness. However, (28) focused on older adults with depression and included the Internet and computers only as technological interventions whereas we included all types of digital technology interventions and adults of all age groups (≥ 18 years). In addition, studies (n=5) included in the meta-analysis by (28) had different follow-up periods (3-6 months) but they did not report which follow-up measurements they included in their meta-analysis. We ran different meta-analyses for measurements at different follow-up periods, i.e. three, four and six months, as suggested by the Cochrane guidelines (47).

In addition, a meta-analysis by Masi et al., 2011 also reported that technological interventions reduce loneliness, which was more in pre-post studies and non-randomised studies compared to RCTs. However, they included studies with technology and non-technology based interventions (25) whereas we focused on studies with DTIs only. Masi et al., 2011 did not report how they used measurements at different follow-up periods while we avoided combining follow-up measurements at different times as suggested (47). Nonetheless, Masi et al concluded that the technology is yet to be capitalised on for loneliness (25), which is evident from our findings that show that DTIs do not reduce loneliness, especially in adults.

Interestingly, our findings provide new insights about loneliness in older adults. Despite our inclusion criteria of age (18 years and above), the selected studies more commonly involved older people (averagely 70+ years old) and there was no statistically significant reduction in loneliness. Our findings might suggest that DTIs do not reduce loneliness in adults, which is opposite to a commonly held view that digital technology can solve the problem of loneliness, especially in older people. However, the digital technologies are tools and means to social connectedness, which may help in lessening loneliness for a little while because the effects of DTIs are short-lived and do not last beyond six months of the intervention (66). In addition, digital technologies might provide digital social connection but in fact they reduce social connectedness in real life (67). Digital technologies are only tools that extend opportunities for connecting with others but do not provide real human interaction (68) and cannot replace human contact (40).

### 4.2 Limitations

Limitations of our systematic reviews include a small number of studies (n=6) with heterogeneous sets of results, the selection of English language peer reviewed literature published from January 1, 2010 to July 31, 2019 and the intervention duration minimum of three months, which could have resulted in the inclusion of a low number of studies and possible exclusion of potential studies that would have provided useful evidence.

In addition, we could not run sub-group and meta-regression analyses due to very limited number of studies and lack of data on loneliness by participants’ demographic characteristics. In addition, our review might be narrow because we excluded some studies (39, 69-75), which met the technology criterion such as the use of robots, sensors, digital speakers and apps but failed to meet other selection criteria. Thus, our meta-analysis was limited to studies mostly about social interactions using digital tools.

Moreover, another limitation could be using a meta-analysis based on only the follow-up data. For example, study by Tsai et al., 2010 in the 3 months follow-up meta-analysis had an SMD of 0.06 with a CI −0.8 to +0.65 (p=0.03) (Fig. 2), which may suggest that these studies may have had higher power to show a difference compared to baseline loneliness.

As recommendations for future research, we suggest that researchers involved in trials may agree on a common measure of loneliness and consider reporting of results in a standardised way that would allow pooling of baseline-adjusted estimates of the treatment effect rather than differences in follow-up means.

## 5 CONCLUSIONS

We did not find sufficient evidence supporting the effectiveness of digital technology intervention to reduce loneliness in (older) adults. Our findings may suggest further research involving RCTs (44) with larger sample sizes with longer duration of interventions and follow-up measurement periods. Future research may apply inclusive research designs using a combination of digital applications including robots, sensors and social connecting applications by involving adults in the age group 45-65 years as this segment of the population is more likely to be more technology savvy and digital interventions might be more effective. Future research might also target ethnic, racial, and sexual orientation minority communities where loneliness is common (11).

## Data Availability

All data are reported in the manuscript.

## Previous presentation

A poster on the preliminary findings of our systematic review presented at the 12th European Public Health Conference, 20–23 November 2019, Marseille, France. The abstract of the poster was published in the European Journal of Public Health, Vol. 29, Supplement 4, 2019

## Acknowledgements

The authors acknowledge support from Liz Callow of Bodleian Health Care Libraries, University of Oxford for running literature searches. We gratefully thank to Dr Alexandra Farrow, Brunel University London for checking the manuscript.

## Appendix 1. Literature searches

| Search Query | DATABASES SEARCHED |  |  |  |  |
| --- | --- | --- | --- | --- | --- |
|  | PubMed | EMBASE | Medline | CINAHL | Web of Science |
| **<u>Subject Headings</u>** |  | *(via OVID)* | *(via OVID)* | *(via EBSCOHOST)* |  |
| Loneliness | Yes | Yes | Yes | Yes | No subject headings |
| Technology | No | No | No | Yes | No subject headings |
| Robotics | Yes | Yes | Yes | Yes | No subject headings |
| Internet | Yes | Yes | Yes | Yes | No subject headings |
| Social Media | Yes | Yes | Yes | Yes | No subject headings |
| Telephone | Yes | Yes | Yes | Yes | No subject headings |
| Smartphone | Yes | Yes | Yes | Yes | No subject headings |
| Computer | Yes | Yes | Yes | Yes | No subject headings |
| Computer handheld | Yes | No | Yes | Yes | No subject headings |
| Personal digital assistant | No | Yes | No | No | No subject headings |
| Web (world Wide Web) | No | No | No | Yes | No subject headings |
| **<u>Phrases / text</u>** |  |  |  |  |  |
| Loneliness **OR** lonely **OR** social isolation [Title/Abstract] | Yes | Yes | Yes | Yes | **Searched as topics** (includes: Title, Abstract, Author Keywords, Keywords Plus) |
| **AND** |  |  |  |  |  |
| Digital* **OR** technolog* **OR** sensor* **OR** robot* **OR** internet **OR** social media **OR** smartphone* **OR** smart phone* **OR** telephone* **OR** phone **OR** online **OR** ipad* **OR** computer* **OR** electronic* **OR** Web [Title/Abstract] | Yes | Yes | Yes | Yes | digital* **OR** technolog* **OR** sensor* **OR** robot* **OR** internet **OR** social media **OR** smartphone* **OR** online **OR** ipad* **OR** computer* **OR** electronic* **OR** Web [**Searched as topics**] |
| **<u>Filters</u>** |  |  |  |  |  |
| **Publication years:** 2010-2019 | Yes | Yes | Yes | Yes | Yes |
| **Language:** English | Yes | Yes | Yes | Yes | Yes |
| **Species:** Humans | Yes | Yes | Yes | Yes | N/A |
| **Document types:** | Clinical Trial,<br>Controlled<br>Clinical Trial,<br>Evaluation<br>Studies,<br>Journal<br>Article,<br>Meta-Analysis,<br>Observational<br>Study,<br>Randomized<br>Controlled<br>Trial,<br>Review,<br>Systematic<br>Reviews | Trials (all) | Clinical Trial,<br>Evaluation Studies<br>Journal Article<br>Meta-Analysis<br>Observational<br>Study<br>Review<br>Systematic<br>Reviews | Clinical Trial,<br>Journal Article,<br>Meta-Analysis,<br>Randomized<br>Controlled Trial,<br>Research, Review<br>Systematic Review | Articles, Reviews |

### PUBMED SEARCHES

((“Loneliness”[Mesh]) OR (((loneliness[Title/Abstract]) OR lonely[Title/Abstract]) OR social isolation[Title/Abstract])) AND ((“Robotics”[Mesh]) OR (“Internet”[Mesh]) OR (“Social Media”[Mesh]) OR (“Smartphone”[Mesh]) OR (“Telephone”[Mesh]) OR (“Computers, Handheld”[Mesh]) OR (“Computers”[Mesh]) OR (((((((((((((digital*[Title/Abstract]) OR technolog*[Title/Abstract]) OR sensor*[Title/Abstract]) OR robot*[Title/Abstract]) OR internet[Title/Abstract]) OR social media[Title/Abstract]) OR phone*[Title/Abstract]) OR telephone*[Title/Abstract]) OR online[Title/Abstract]) OR ipad*[Title/Abstract]) OR computer*[Title/Abstract]) OR electronic*[Title/Abstract]) OR web[Title/Abstract]))

(“Loneliness”[Mesh] OR ((loneliness[Title/Abstract] OR lonely[Title/Abstract]) OR social isolation[Title/Abstract])) AND (“Robotics”[Mesh] OR “Internet”[Mesh] OR “Social Media”[Mesh] OR “Smartphone”[Mesh] OR “Telephone”[Mesh] OR “Computers, Handheld”[Mesh] OR “Computers”[Mesh] OR (((((((((((((digital[Title/Abstract] OR digital“[Title/Abstract] OR digital”[Title/Abstract] OR digital’s[Title/Abstract] OR digital1[Title/Abstract] OR digitala[Title/Abstract] OR digitalassets[Title/Abstract] OR digitalb[Title/Abstract] OR digitalcell[Title/Abstract] OR digitalcellsorter[Title/Abstract] OR digitalcommunication[Title/Abstract] OR digitaldiagnost[Title/Abstract] OR digitaldlsorter[Title/Abstract] OR digitaldna[Title/Abstract] OR digitale[Title/Abstract] OR digitaleae[Title/Abstract] OR digitalel[Title/Abstract] OR digitalemia[Title/Abstract] OR digitalemia’s[Title/Abstract] OR digitalemic[Title/Abstract] OR digitalen[Title/Abstract] OR digitaler[Title/Abstract] OR digitalera[Title/Abstract] OR digitales[Title/Abstract] OR digitalface[Title/Abstract] OR digitalfiltering[Title/Abstract] OR digitalfishlibrary[Title/Abstract] OR digitalgia[Title/Abstract] OR digitalglobe[Title/Abstract] OR digitalhealth[Title/Abstract] OR digitalhealtheurope[Title/Abstract] OR digitalhealthscore[Title/Abstract] OR digitalhub[Title/Abstract] OR digitalhub’s[Title/Abstract] OR digitali[Title/Abstract] OR digitalia[Title/Abstract] OR digitalic[Title/Abstract] OR digitalica[Title/Abstract] OR digitalics[Title/Abstract] OR digitalideae[Title/Abstract] OR digitalidis[Title/Abstract] OR digitalin[Title/Abstract] OR digitalinaemia[Title/Abstract] OR digitaline[Title/Abstract] OR digitalins[Title/Abstract] OR digitalinum[Title/Abstract] OR digitalis[Title/Abstract] OR digitalis’[Title/Abstract] OR digitalis’s[Title/Abstract] OR digitalisation[Title/Abstract] OR digitalisconcentration[Title/Abstract] OR digitalisconcentrations[Title/Abstract] OR digitalise[Title/Abstract] OR digitalised[Title/Abstract] OR digitalisglycoside[Title/Abstract] OR digitalisglycosides[Title/Abstract] OR digitalisierten[Title/Abstract] OR digitalisierung[Title/Abstract] OR digitalising[Title/Abstract] OR digitalisintoxication[Title/Abstract] OR digitalisintoxications[Title/Abstract] OR digitalisize[Title/Abstract] OR digitalislike[Title/Abstract] OR digitalism[Title/Abstract] OR digitalispreparations[Title/Abstract] OR digitalisreceptor[Title/Abstract] OR digitality[Title/Abstract] OR digitalizacion[Title/Abstract] OR digitalizada[Title/Abstract] OR digitalizadas[Title/Abstract] OR digitalizaing[Title/Abstract] OR digitalization[Title/Abstract] OR digitalization’[Title/Abstract] OR digitalizations[Title/Abstract] OR digitalize[Title/Abstract] OR digitalized[Title/Abstract] OR digitalizer[Title/Abstract] OR digitalizers[Title/Abstract] OR digitalizes[Title/Abstract] OR digitalizing[Title/Abstract] OR digitalizzata[Title/Abstract] OR digitalizzazione[Title/Abstract] OR digitaljournal[Title/Abstract] OR digitalk[Title/Abstract] OR digitall[Title/Abstract] OR digitallis[Title/Abstract] OR digitallung[Title/Abstract] OR digitally[Title/Abstract] OR digitally’[Title/Abstract] OR digitallyconnected[Title/Abstract] OR digitalmammography[Title/Abstract] OR digitalme[Title/Abstract] OR digitalmed[Title/Abstract] OR digitalmetrade[Title/Abstract] OR digitalmicrograph[Title/Abstract] OR digitalmicrographtrade[Title/Abstract] OR digitalmill[Title/Abstract] OR digitalmlpa[Title/Abstract] OR digitalo[Title/Abstract] OR digitaloid[Title/Abstract] OR digitaloides[Title/Abstract] OR digitaloidites[Title/Abstract] OR digitaloids[Title/Abstract] OR digitalonin[Title/Abstract] OR digitalopyranoside[Title/Abstract] OR digitalopyranosyl[Title/Abstract] OR digitalose[Title/Abstract] OR digitalosid[Title/Abstract] OR digitaloside[Title/Abstract] OR digitalosyl[Title/Abstract] OR digitalotherapy[Title/Abstract] OR digitalpcr[Title/Abstract] OR digitalpt[Title/Abstract] OR digitalradiography[Title/Abstract] OR digitalrectal[Title/Abstract] OR digitalrom[Title/Abstract] OR digitals[Title/Abstract] OR digitalslide[Title/Abstract] OR digitalslidearchive[Title/Abstract] OR digitalspiders[Title/Abstract] OR digitaltf[Title/Abstract] OR digitaltrade[Title/Abstract] OR digitalvhi[Title/Abstract] OR digitalyzed[Title/Abstract]) OR (technolog[Title/Abstract] OR technologeous[Title/Abstract] OR technologia[Title/Abstract] OR technologiae[Title/Abstract] OR technologic[Title/Abstract] OR technological[Title/Abstract] OR technological’[Title/Abstract] OR technologicalchallenge[Title/Abstract] OR technologicalforecasting[Title/Abstract] OR technologicalforesight[Title/Abstract] OR technologicalization[Title/Abstract] OR technologically[Title/Abstract] OR technologically’[Title/Abstract] OR technologicalperspective[Title/Abstract] OR technologics[Title/Abstract] OR technologicus[Title/Abstract] OR technologie[Title/Abstract] OR technologie’[Title/Abstract] OR technologiebewertung[Title/Abstract] OR technologiees[Title/Abstract] OR technologielaan[Title/Abstract] OR technologien[Title/Abstract] OR technologiepark[Title/Abstract] OR technologies[Title/Abstract] OR technologies’[Title/Abstract] OR technologies’s[Title/Abstract] OR technologiesare[Title/Abstract] OR technologiesartificial[Title/Abstract] OR technologiesguidance[Title/Abstract] OR technologiesis[Title/Abstract] OR technologiesrome[Title/Abstract] OR technologiessuch[Title/Abstract] OR technologiestm[Title/Abstract] OR technologiestrade[Title/Abstract] OR technologieswe[Title/Abstract] OR technologii[Title/Abstract] OR technologique[Title/Abstract] OR technologiques[Title/Abstract] OR technologis[Title/Abstract] OR technologisation[Title/Abstract] OR technologische[Title/Abstract] OR technologised[Title/Abstract] OR technologising[Title/Abstract] OR technologism[Title/Abstract] OR technologist[Title/Abstract] OR technologist’[Title/Abstract] OR technologist’s[Title/Abstract] OR technologists[Title/Abstract] OR technologists’[Title/Abstract] OR technologists’attitudes[Title/Abstract] OR technologists’s[Title/Abstract] OR technologization[Title/Abstract] OR technologize[Title/Abstract] OR technologized[Title/Abstract] OR technologizing[Title/Abstract] OR technologlans[Title/Abstract] OR technologly[Title/Abstract] OR technology[Title/Abstract] OR technology’[Title/Abstract] OR technology’’[Title/Abstract] OR technology’s[Title/Abstract] OR technology1[Title/Abstract] OR technology16[Title/Abstract] OR technology3[Title/Abstract] OR technologyallows[Title/Abstract] OR technologyand[Title/Abstract] OR technologyassisted[Title/Abstract] OR technologyconsider[Title/Abstract] OR technologydagger[Title/Abstract] OR technologyevaluation[Title/Abstract] OR technologyfor[Title/Abstract] OR technologyhave[Title/Abstract] OR technologyhose[Title/Abstract] OR technologyin[Title/Abstract] OR technologyis[Title/Abstract] OR technologyit[Title/Abstract] OR technologymc[Title/Abstract] OR technologypreclinical[Title/Abstract] OR technologys[Title/Abstract] OR technologythat[Title/Abstract] OR technologyto[Title/Abstract] OR technologytrade[Title/Abstract] OR technologytranslation[Title/Abstract] OR technologyuser[Title/Abstract] OR technologyvarious[Title/Abstract])) OR (sensor[Title/Abstract] OR sensor’[Title/Abstract] OR sensor’s[Title/Abstract] OR sensor1[Title/Abstract] OR sensor4pri[Title/Abstract] OR sensora[Title/Abstract] OR sensorad[Title/Abstract] OR sensoraid[Title/Abstract] OR sensoral[Title/Abstract] OR sensorale[Title/Abstract] OR sensorally[Title/Abstract] OR sensorand[Title/Abstract] OR sensorant[Title/Abstract] OR sensorapertures[Title/Abstract] OR sensorarray[Title/Abstract] OR sensorart[Title/Abstract] OR sensorband[Title/Abstract] OR sensorbility[Title/Abstract] OR sensorbod[Title/Abstract] OR sensorbox[Title/Abstract] OR sensorcaine[Title/Abstract] OR sensorcatheter[Title/Abstract] OR sensorchip[Title/Abstract] OR sensorchips[Title/Abstract] OR sensorcm[Title/Abstract] OR sensorconsists[Title/Abstract] OR sensordata[Title/Abstract] OR sensordb[Title/Abstract] OR sensordish[Title/Abstract] OR sensore[Title/Abstract] OR sensoready[Title/Abstract] OR sensorealization[Title/Abstract] OR sensored[Title/Abstract] OR sensoredge[Title/Abstract] OR sensoregulator[Title/Abstract] OR sensoremoval[Title/Abstract] OR sensoremoval’[Title/Abstract] OR sensores[Title/Abstract] OR sensorevery[Title/Abstract] OR sensoreverywhere[Title/Abstract] OR sensorex[Title/Abstract] OR sensorexhibited[Title/Abstract] OR sensorfor[Title/Abstract] OR sensorfret[Title/Abstract] OR sensorg[Title/Abstract] OR sensorgraft[Title/Abstract] OR sensorgram[Title/Abstract] OR sensorgrams[Title/Abstract] OR sensorgraphy[Title/Abstract] OR sensorhis[Title/Abstract] OR sensori[Title/Abstract] OR sensoria[Title/Abstract] OR sensoriactuators[Title/Abstract] OR sensoriais[Title/Abstract] OR sensorial[Title/Abstract] OR sensorial’[Title/Abstract] OR sensoriales[Title/Abstract] OR sensorialist[Title/Abstract] OR sensorialite[Title/Abstract] OR sensoriality[Title/Abstract] OR sensorialized[Title/Abstract] OR sensorially[Title/Abstract] OR sensorials[Title/Abstract] OR sensorialy[Title/Abstract] OR sensoribehavioural[Title/Abstract] OR sensoric[Title/Abstract] OR sensorica[Title/Abstract] OR sensorical[Title/Abstract] OR sensorically[Title/Abstract] OR sensoricortical[Title/Abstract] OR sensoricphysiological[Title/Abstract] OR sensorics[Title/Abstract] OR sensoridiscriminative[Title/Abstract] OR sensoriel[Title/Abstract] OR sensorielle[Title/Abstract] OR sensorielles[Title/Abstract] OR sensoriels[Title/Abstract] OR sensories[Title/Abstract] OR sensorif[Title/Abstract] OR sensorigenesis[Title/Abstract] OR sensorii[Title/Abstract] OR sensoril[Title/Abstract] OR sensorilimbic[Title/Abstract] OR sensorily[Title/Abstract] OR sensorimetry[Title/Abstract] OR sensorimortor[Title/Abstract] OR sensorimoter[Title/Abstract] OR sensorimoteur[Title/Abstract] OR sensorimoteurs[Title/Abstract] OR sensorimotility[Title/Abstract] OR sensorimotor[Title/Abstract] OR sensorimotor’[Title/Abstract] OR sensorimotoraffective[Title/Abstract] OR sensorimotoraxonal[Title/Abstract] OR sensorimotorcontrol[Title/Abstract] OR sensorimotorcortical[Title/Abstract] OR sensorimotori[Title/Abstract] OR sensorimotorial[Title/Abstract] OR sensorimotoric[Title/Abstract] OR sensorimotorrhythm[Title/Abstract] OR sensorimotorsystem[Title/Abstract] OR sensorimotory[Title/Abstract] OR sensorimototor[Title/Abstract] OR sensorimotrice[Title/Abstract] OR sensorimotrices[Title/Abstract] OR sensorimotricity[Title/Abstract] OR sensorin[Title/Abstract] OR sensorincludes[Title/Abstract] OR sensorineal[Title/Abstract] OR sensorineral[Title/Abstract] OR sensorineraul[Title/Abstract] OR sensorinerual[Title/Abstract] OR sensorinerural[Title/Abstract] OR sensoriness[Title/Abstract] OR sensorineu[Title/Abstract] OR sensorineual[Title/Abstract] OR sensorineueal[Title/Abstract] OR sensorineumral[Title/Abstract] OR sensorineur[Title/Abstract] OR sensorineura[Title/Abstract] OR sensorineural[Title/Abstract] OR sensorineural’[Title/Abstract] OR sensorineuralhearing[Title/Abstract] OR sensorineuralis[Title/Abstract] OR sensorineurally[Title/Abstract] OR sensorineurals[Title/Abstract] OR sensorineurepithelium[Title/Abstract] OR sensorineurinal[Title/Abstract] OR sensorineurocognitive[Title/Abstract] OR sensorineuroepithelium[Title/Abstract] OR sensorineurological[Title/Abstract] OR sensorineuronal[Title/Abstract] OR sensorineutral[Title/Abstract] OR sensoring[Title/Abstract] OR sensoringeural[Title/Abstract] OR sensorintegrative[Title/Abstract] OR sensorinueral[Title/Abstract] OR sensorinural[Title/Abstract] OR sensorio[Title/Abstract] OR sensorioculomotor[Title/Abstract] OR sensoriomotor[Title/Abstract] OR sensoriomotora[Title/Abstract] OR sensoriomotoras[Title/Abstract] OR sensoriomotriz[Title/Abstract] OR sensorion[Title/Abstract] OR sensorioneural[Title/Abstract] OR sensoriperception[Title/Abstract] OR sensoriperceptual[Title/Abstract] OR sensoriphasic[Title/Abstract] OR sensoriphobia[Title/Abstract] OR sensoriprocessing[Title/Abstract] OR sensoris[Title/Abstract] OR sensorisation[Title/Abstract] OR sensorische[Title/Abstract] OR sensorised[Title/Abstract] OR sensorisensory[Title/Abstract] OR sensoriske[Title/Abstract] OR sensorisomatic[Title/Abstract] OR sensoristasis[Title/Abstract] OR sensoristatic[Title/Abstract] OR sensoristic[Title/Abstract] OR sensoristics[Title/Abstract] OR sensoristrain[Title/Abstract] OR sensoristrain’[Title/Abstract] OR sensorithm[Title/Abstract] OR sensoritmotor[Title/Abstract] OR sensoritopic[Title/Abstract] OR sensoritrigeminal[Title/Abstract] OR sensoritrophic[Title/Abstract] OR sensority[Title/Abstract] OR sensorium[Title/Abstract] OR sensorium’[Title/Abstract] OR sensoriums[Title/Abstract] OR sensorius[Title/Abstract] OR sensorivagal[Title/Abstract] OR sensorization[Title/Abstract] OR sensorize[Title/Abstract] OR sensorized[Title/Abstract] OR sensorized’[Title/Abstract] OR sensorizing[Title/Abstract] OR sensorless[Title/Abstract] OR sensorlike[Title/Abstract] OR sensorlink[Title/Abstract] OR sensorlog[Title/Abstract] OR sensormat[Title/Abstract] OR sensormedic[Title/Abstract] OR sensormedics[Title/Abstract] OR sensormedicus[Title/Abstract] OR sensormedix[Title/Abstract] OR sensorml[Title/Abstract] OR sensormotor[Title/Abstract] OR sensornet[Title/Abstract] OR sensornetworks[Title/Abstract] OR sensorneural[Title/Abstract] OR sensornodes[Title/Abstract] OR sensornye[Title/Abstract] OR sensoro[Title/Abstract] OR sensorograms[Title/Abstract] OR sensoromotor[Title/Abstract] OR sensoromotoric[Title/Abstract] OR sensoroneural[Title/Abstract] OR sensoror[Title/Abstract] OR sensorotoxin[Title/Abstract] OR sensorpoly[Title/Abstract] OR sensorproperties[Title/Abstract] OR sensorregulator[Title/Abstract] OR sensorreporters[Title/Abstract] OR sensorresponse[Title/Abstract] OR sensors[Title/Abstract] OR sensors’[Title/Abstract] OR sensors’s[Title/Abstract] OR sensors2016[Title/Abstract] OR sensorsa[Title/Abstract] OR sensorsand[Title/Abstract] OR sensorsbased[Title/Abstract] OR sensorscope[Title/Abstract] OR sensorselection[Title/Abstract] OR sensorsetting[Title/Abstract] OR sensorship[Title/Abstract] OR sensorshowed[Title/Abstract] OR sensorsi[Title/Abstract] OR sensorsis[Title/Abstract] OR sensorspace[Title/Abstract] OR sensorsthe[Title/Abstract] OR sensorstrips[Title/Abstract] OR sensorsuse[Title/Abstract] OR sensorsv[Title/Abstract] OR sensorswere[Title/Abstract] OR sensort[Title/Abstract] OR sensortag[Title/Abstract] OR sensortalk[Title/Abstract] OR sensortech[Title/Abstract] OR sensortechnik[Title/Abstract] OR sensortechnology[Title/Abstract] OR sensortek[Title/Abstract] OR sensorthat[Title/Abstract] OR sensorthings[Title/Abstract] OR sensorto[Title/Abstract] OR sensortouch[Title/Abstract] OR sensortrade[Title/Abstract] OR sensorts[Title/Abstract] OR sensorwas[Title/Abstract] OR sensorweb[Title/Abstract] OR sensory[Title/Abstract] OR sensory’[Title/Abstract] OR sensory’s[Title/Abstract] OR sensoryanalysis[Title/Abstract] OR sensorydeprivation[Title/Abstract] OR sensoryevoked[Title/Abstract] OR sensorygc[Title/Abstract] OR sensoryical[Title/Abstract] OR sensoryinteractions[Title/Abstract] OR sensorylike[Title/Abstract] OR sensorymodality[Title/Abstract] OR sensorymotor[Title/Abstract] OR sensorynav1[Title/Abstract] OR sensoryneural[Title/Abstract] OR sensoryneuropathy[Title/Abstract] OR sensoryomics[Title/Abstract] OR sensorypredominant[Title/Abstract] OR sensoryprocessing[Title/Abstract] OR sensoryrecovery[Title/Abstract] OR sensoryrelated[Title/Abstract] OR sensoryrhodopsin[Title/Abstract] OR sensoryrizotomy[Title/Abstract] OR sensorytransduction[Title/Abstract] OR sensorytreat[Title/Abstract] OR sensorytreat’s[Title/Abstract])) OR (robot[Title/Abstract] OR robot’[Title/Abstract] OR robot’consists[Title/Abstract] OR robot’electronically[Title/Abstract] OR robot’s[Title/Abstract] OR robota[Title/Abstract] OR robotac[Title/Abstract] OR robotanalyst[Title/Abstract] OR robotanalyst’s[Title/Abstract] OR robotarium[Title/Abstract] OR robotassisted[Title/Abstract] OR robotassistedlaparoscopic[Title/Abstract] OR robotcar[Title/Abstract] OR robotechnologies[Title/Abstract] OR robotechnologist[Title/Abstract] OR roboteeg[Title/Abstract] OR roboter[Title/Abstract] OR roboterassistiert[Title/Abstract] OR roboterassistierte[Title/Abstract] OR roboterassistierter[Title/Abstract] OR robotfoto[Title/Abstract] OR robotham[Title/Abstract] OR robotherapist[Title/Abstract] OR robotherapist’[Title/Abstract] OR robotherapy[Title/Abstract] OR robothics[Title/Abstract] OR roboti[Title/Abstract] OR robotic[Title/Abstract] OR robotic’[Title/Abstract] OR robotica[Title/Abstract] OR robotical[Title/Abstract] OR robotically[Title/Abstract] OR roboticallyassisted[Title/Abstract] OR roboticamente[Title/Abstract] OR roboticapproach[Title/Abstract] OR roboticarm[Title/Abstract] OR roboticas[Title/Abstract] OR roboticassisted[Title/Abstract] OR roboticbed[Title/Abstract] OR roboticbird[Title/Abstract] OR roboticians[Title/Abstract] OR roboticist[Title/Abstract] OR roboticists[Title/Abstract] OR roboticists’[Title/Abstract] OR roboticization[Title/Abstract] OR roboticized[Title/Abstract] OR roboticizes[Title/Abstract] OR robotico[Title/Abstract] OR roboticos[Title/Abstract] OR roboticpancreatoduodenectomy[Title/Abstract] OR robotics[Title/Abstract] OR robotics’[Title/Abstract] OR roboticslab[Title/Abstract] OR roboticsurgery[Title/Abstract] OR roboticsystem[Title/Abstract] OR robotik[Title/Abstract] OR robotiker[Title/Abstract] OR robotiq[Title/Abstract] OR robotique[Title/Abstract] OR robotiquette[Title/Abstract] OR robotiquette’[Title/Abstract] OR robotisation[Title/Abstract] OR robotisches[Title/Abstract] OR robotise[Title/Abstract] OR robotised[Title/Abstract] OR robotisee[Title/Abstract] OR robotitian[Title/Abstract] OR robotix[Title/Abstract] OR robotization[Title/Abstract] OR robotize[Title/Abstract] OR robotized[Title/Abstract] OR robotizing[Title/Abstract] OR robotizzato[Title/Abstract] OR robotless[Title/Abstract] OR robotlike[Title/Abstract] OR robotmajsebeszet[Title/Abstract] OR robotmanipulators[Title/Abstract] OR robotmediated[Title/Abstract] OR robotnikinin[Title/Abstract] OR robotnon[Title/Abstract] OR roboto[Title/Abstract] OR robotocs[Title/Abstract] OR robotok[Title/Abstract] OR robotol[Title/Abstract] OR robototherapy[Title/Abstract] OR robotrac[Title/Abstract] OR robotrat[Title/Abstract] OR robotreviewer[Title/Abstract] OR robotreviewer’s[Title/Abstract] OR robotripping[Title/Abstract] OR robotrobot[Title/Abstract] OR robotron[Title/Abstract] OR robots[Title/Abstract] OR robots’[Title/Abstract] OR robots’designs[Title/Abstract] OR robotsci[Title/Abstract] OR robotscientist[Title/Abstract] OR robotsebeszet[Title/Abstract] OR robotsebeszeti[Title/Abstract] OR robotsfor[Title/Abstract] OR robotti[Title/Abstract] OR robottom[Title/Abstract] OR robottrade[Title/Abstract] OR robotuna[Title/Abstract] OR robotutor[Title/Abstract] OR robotworld[Title/Abstract])) OR internet[Title/Abstract]) OR social media[Title/Abstract]) OR (phone[Title/Abstract] OR phone’[Title/Abstract] OR phone’s[Title/Abstract] OR phonear[Title/Abstract] OR phonebased[Title/Abstract] OR phonebook[Title/Abstract] OR phoneburst[Title/Abstract] OR phonecall[Title/Abstract] OR phonecard[Title/Abstract] OR phonecardiographic[Title/Abstract] OR phonecians[Title/Abstract] OR phonecians’[Title/Abstract] OR phonectically[Title/Abstract] OR phoned[Title/Abstract] OR phonedependency[Title/Abstract] OR phoneeded[Title/Abstract] OR phoneeutria[Title/Abstract] OR phonefriend[Title/Abstract] OR phonegap[Title/Abstract] OR phoneigen[Title/Abstract] OR phonein[Title/Abstract] OR phoneix[Title/Abstract] OR phonelectrocardiography[Title/Abstract] OR phoneline[Title/Abstract] OR phonem[Title/Abstract] OR phonema[Title/Abstract] OR phonemas[Title/Abstract] OR phonematic[Title/Abstract] OR phonematically[Title/Abstract] OR phonematics[Title/Abstract] OR phonematique[Title/Abstract] OR phonemchanograms[Title/Abstract] OR phoneme[Title/Abstract] OR phoneme’[Title/Abstract] OR phoneme’s[Title/Abstract] OR phonemegrapheme[Title/Abstract] OR phonemena[Title/Abstract] OR phonemes[Title/Abstract] OR phonemes’[Title/Abstract] OR phonemeter[Title/Abstract] OR phonemethod[Title/Abstract] OR phonemic[Title/Abstract] OR phonemicably[Title/Abstract] OR phonemically[Title/Abstract] OR phonemicisation[Title/Abstract] OR phonemicization[Title/Abstract] OR phonemics[Title/Abstract] OR phonemisation[Title/Abstract] OR phonemoformation[Title/Abstract] OR phonemonon[Title/Abstract] OR phonemotopic[Title/Abstract] OR phonems[Title/Abstract] OR phonendoscope[Title/Abstract] OR phonendoscopes[Title/Abstract] OR phonendoscopic[Title/Abstract] OR phonenix[Title/Abstract] OR phonenterographia[Title/Abstract] OR phoneomic[Title/Abstract] OR phonequant[Title/Abstract] OR phonequit[Title/Abstract] OR phonereporting[Title/Abstract] OR phonernically[Title/Abstract] OR phonerpeton[Title/Abstract] OR phones[Title/Abstract] OR phones’[Title/Abstract] OR phones4u[Title/Abstract] OR phoneshop[Title/Abstract] OR phoneside[Title/Abstract] OR phonesimulated[Title/Abstract] OR phonesis[Title/Abstract] OR phonesoap[Title/Abstract] OR phonesonhuman[Title/Abstract] OR phonesthemes[Title/Abstract] OR phonesthemic[Title/Abstract] OR phonet[Title/Abstract] OR phonethep[Title/Abstract] OR phonetic[Title/Abstract] OR phonetica[Title/Abstract] OR phonetical[Title/Abstract] OR phonetically[Title/Abstract] OR phoneticeffects[Title/Abstract] OR phonetician[Title/Abstract] OR phonetician’s[Title/Abstract] OR phoneticians[Title/Abstract] OR phoneticization[Title/Abstract] OR phoneticized[Title/Abstract] OR phonetics[Title/Abstract] OR phonetics’[Title/Abstract] OR phonetik[Title/Abstract] OR phonetion[Title/Abstract] OR phonetique[Title/Abstract] OR phonetisation[Title/Abstract] OR phonetogram[Title/Abstract] OR phonetograms[Title/Abstract] OR phonetograph[Title/Abstract] OR phonetographic[Title/Abstract] OR phonetography[Title/Abstract] OR phonetometric[Title/Abstract] OR phonetoxin[Title/Abstract] OR phonetype[Title/Abstract] OR phonetypical[Title/Abstract] OR phoneus[Title/Abstract] OR phoneuse[Title/Abstract] OR phoneutria[Title/Abstract] OR phoneutriatoxin[Title/Abstract] OR phoneutrism[Title/Abstract] OR phoney[Title/Abstract] OR phoneyusa[Title/Abstract])) OR (telephone[Title/Abstract] OR telephone’[Title/Abstract] OR telephone’s[Title/Abstract] OR telephoneassisted[Title/Abstract] OR telephonecare[Title/Abstract] OR telephoned[Title/Abstract] OR telephoneear[Title/Abstract] OR telephoneinterviews[Title/Abstract] OR telephoners[Title/Abstract] OR telephoners’[Title/Abstract] OR telephones[Title/Abstract] OR telephonesex[Title/Abstract] OR telephonetics[Title/Abstract])) OR online[Title/Abstract]) OR (ipad[Title/Abstract] OR ipad’s[Title/Abstract] OR ipad120[Title/Abstract] OR ipad2[Title/Abstract] OR ipad20[Title/Abstract] OR ipad2s[Title/Abstract] OR ipad4[Title/Abstract] OR ipada[Title/Abstract] OR ipadair[Title/Abstract] OR ipadam[Title/Abstract] OR ipadar[Title/Abstract] OR ipadb[Title/Abstract] OR ipade[Title/Abstract] OR ipadech[Title/Abstract] OR ipadh[Title/Abstract] OR ipadia[Title/Abstract] OR ipadm[Title/Abstract] OR ipadnych[Title/Abstract] OR ipado[Title/Abstract] OR ipados[Title/Abstract] OR ipads[Title/Abstract] OR ipadstrade[Title/Abstract] OR ipadt[Title/Abstract] OR ipadtrade[Title/Abstract] OR ipadu[Title/Abstract] OR ipadur1[Title/Abstract] OR ipadvas[Title/Abstract])) OR (computer[Title/Abstract] OR computer’[Title/Abstract] OR computer’s[Title/Abstract] OR computer18[Title/Abstract] OR computer19[Title/Abstract] OR computer3[Title/Abstract] OR computeradapted[Title/Abstract] OR computeraided[Title/Abstract] OR computerally[Title/Abstract] OR computeranalysis[Title/Abstract] OR computerand[Title/Abstract] OR computerangiography[Title/Abstract] OR computeranimated[Title/Abstract] OR computerarthrometry[Title/Abstract] OR computerassisted[Title/Abstract] OR computerassistierte[Title/Abstract] OR computerbase[Title/Abstract] OR computerbased[Title/Abstract] OR computercalculated[Title/Abstract] OR computerchemie[Title/Abstract] OR computerclassified[Title/Abstract] OR computercode[Title/Abstract] OR computercontrolled[Title/Abstract] OR computerd[Title/Abstract] OR computerdesigned[Title/Abstract] OR computerdocking[Title/Abstract] OR computerdokumentation[Title/Abstract] OR computerdriven[Title/Abstract] OR computered[Title/Abstract] OR computerenhanced[Title/Abstract] OR computerese[Title/Abstract] OR computereyes[Title/Abstract] OR computerezed[Title/Abstract] OR computerfiled[Title/Abstract] OR computerfitted[Title/Abstract] OR computergame[Title/Abstract] OR computergames[Title/Abstract] OR computergenerated[Title/Abstract] OR computergestutzte[Title/Abstract] OR computergestutzter[Title/Abstract] OR computergraphic[Title/Abstract] OR computergraphical[Title/Abstract] OR computergraphically[Title/Abstract] OR computergraphics[Title/Abstract] OR computergraphs[Title/Abstract] OR computerguided[Title/Abstract] OR computerheld[Title/Abstract] OR computeric[Title/Abstract] OR computerin[Title/Abstract] OR computering[Title/Abstract] OR computerintensive[Title/Abstract] OR computerinterpolated[Title/Abstract] OR computeris[Title/Abstract] OR computerisation[Title/Abstract] OR computerisations[Title/Abstract] OR computerise[Title/Abstract] OR computerised[Title/Abstract] OR computerised’[Title/Abstract] OR computerisedprovider[Title/Abstract] OR computerisee[Title/Abstract] OR computerises[Title/Abstract] OR computerising[Title/Abstract] OR computerism[Title/Abstract] OR computerizability[Title/Abstract] OR computerizable[Title/Abstract] OR computerizada[Title/Abstract] OR computerization[Title/Abstract] OR computerization’[Title/Abstract] OR computerization’s[Title/Abstract] OR computerizd[Title/Abstract] OR computerize[Title/Abstract] OR computerizea[Title/Abstract] OR computerized[Title/Abstract] OR computerized’[Title/Abstract] OR computerizedcardiotocography[Title/Abstract] OR computerizeddynamic[Title/Abstract] OR computerizeds[Title/Abstract] OR computerizedt[Title/Abstract] OR computerizedtomographic[Title/Abstract] OR computerizes[Title/Abstract] OR computerizewd[Title/Abstract] OR computerizied[Title/Abstract] OR computerizing[Title/Abstract] OR computerizsed[Title/Abstract] OR computerizzata[Title/Abstract] OR computerlab[Title/Abstract] OR computerlabelled[Title/Abstract] OR computerland[Title/Abstract] OR computerless[Title/Abstract] OR computerlike[Title/Abstract] OR computerlink[Title/Abstract] OR computerlink’s[Title/Abstract] OR computerly[Title/Abstract] OR computermediated[Title/Abstract] OR computermen[Title/Abstract] OR computermotion[Title/Abstract] OR computernavigated[Title/Abstract] OR computernavigation[Title/Abstract] OR computerneuron[Title/Abstract] OR computero[Title/Abstract] OR computerogenic[Title/Abstract] OR computerologist[Title/Abstract] OR computeromography[Title/Abstract] OR computeros[Title/Abstract] OR computerpart[Title/Abstract] OR computerphobe[Title/Abstract] OR computerphobia[Title/Abstract] OR computerphobia’[Title/Abstract] OR computerphobic[Title/Abstract] OR computerprogram[Title/Abstract] OR computerprogrammen[Title/Abstract] OR computerprogrammes[Title/Abstract] OR computerprograms[Title/Abstract] OR computerreadable[Title/Abstract] OR computerrelated[Title/Abstract] OR computerrhea[Title/Abstract] OR computerrised[Title/Abstract] OR computers[Title/Abstract] OR computers’[Title/Abstract] OR computers’’[Title/Abstract] OR computerscan[Title/Abstract] OR computerscreen[Title/Abstract] OR computersimulation[Title/Abstract] OR computersimulations[Title/Abstract] OR computersmartphone[Title/Abstract] OR computersonography[Title/Abstract] OR computerspeak[Title/Abstract] OR computerspiel[Title/Abstract] OR computerspielabhangigkeit[Title/Abstract] OR computerspielsucht[Title/Abstract] OR computerstored[Title/Abstract] OR computersupported[Title/Abstract] OR computersystem[Title/Abstract] OR computersystems[Title/Abstract] OR computertime[Title/Abstract] OR computertized[Title/Abstract] OR computertomografie[Title/Abstract] OR computertomografischen[Title/Abstract] OR computertomogram[Title/Abstract] OR computertomogramm[Title/Abstract] OR computertomogramms[Title/Abstract] OR computertomograms[Title/Abstract] OR computertomograph[Title/Abstract] OR computertomographic[Title/Abstract] OR computertomographical[Title/Abstract] OR computertomographically[Title/Abstract] OR computertomographie[Title/Abstract] OR computertomographies[Title/Abstract] OR computertomographs[Title/Abstract] OR computertomography[Title/Abstract] OR computerunterstutzte[Title/Abstract] OR computerunterstutztes[Title/Abstract] OR computerusage[Title/Abstract] OR computerville[Title/Abstract] OR computervirus[Title/Abstract] OR computervision[Title/Abstract] OR computerworld[Title/Abstract])) OR (electronic[Title/Abstract] OR electronic’[Title/Abstract] OR electronic’s[Title/Abstract] OR electronica[Title/Abstract] OR electronical[Title/Abstract] OR electronicall[Title/Abstract] OR electronically[Title/Abstract] OR electronically’[Title/Abstract] OR electronicallyexcited[Title/Abstract] OR electronicaly[Title/Abstract] OR electronicas[Title/Abstract] OR electronicbibliographic[Title/Abstract] OR electroniccards[Title/Abstract] OR electroniccase[Title/Abstract] OR electronicdatabase[Title/Abstract] OR electronicdocument[Title/Abstract] OR electronicfans[Title/Abstract] OR electroniclly[Title/Abstract] OR electronicmaterials[Title/Abstract] OR electronicmedical[Title/Abstract] OR electronicmedsman[Title/Abstract] OR electronicmessage[Title/Abstract] OR electronico[Title/Abstract] OR electronicos[Title/Abstract] OR electronicpatientrecords[Title/Abstract] OR electronicplatform[Title/Abstract] OR electronicquestionnaire[Title/Abstract] OR electronicreadout[Title/Abstract] OR electronicrecruitment[Title/Abstract] OR electronicrograph[Title/Abstract] OR electronics[Title/Abstract] OR electronics’[Title/Abstract] OR electronicsearch[Title/Abstract] OR electronicssensors[Title/Abstract] OR electronictemplate[Title/Abstract] OR electronicwaste[Title/Abstract] OR electronicwe[Title/Abstract])) OR web[Title/Abstract])) AND ((Clinical Trial[ptyp] OR Controlled Clinical Trial[ptyp] OR Journal Article[ptyp] OR Meta-Analysis[ptyp] OR Observational Study[ptyp] OR Randomized Controlled Trial[ptyp] OR Review[ptyp] OR systematic[sb]) AND (“2010/01/01”[PDAT]: “2019/07/31”[PDAT]) AND “humans”[MeSH Terms])

### EMBASE

1. loneliness/
2. Loneliness.ti,ab.
3. (lonely or social isolation).ti,ab.
4. 1 or 2 or 3
5. (digital* or technol* or sensor* or robot* or internet* or social media or smartphone* or smart phone* or telephone* or phone* or online or ipad* or computer* or electronic* or Web).ti,ab.
6. robotics/
7. Internet/
8. social media/
9. smartphone/
10. personal digital assistant/
11. computer/
12. telephone/
13. 5 or 6 or 7 or 8 or 9 or 10 or 11 or 12
14. 4 and 13
15. limit 14 to (human and english language)
16. limit 15 to yr=“2010 -Current”

### MEDLINE

1. LONELINESS/
2. Loneliness.ti,ab.
3. (lonely or social isolation).ti,ab.
4. 1 or 2 or 3
5. (digital* or technol* or sensor* or robot* or internet* or social media or smartphone* or smart phone* or telephone* or phone* or online or ipad* or computer* or electronic* or Web).ti,ab.
6. ROBOTICS/
7. INTERNET/
8. Social Media/
9. TELEPHONE/
10. Smartphone/
11. Computers, Handheld/
12. COMPUTERS/
13. 5 or 6 or 7 or 8 or 9 or 10 or 11 or 12
14. 4 and 13
15. limit 14 to (english language and humans and yr=“2010 -Current” and (clinical trial, all or evaluation studies or journal article or meta analysis or observational study or “review” or systematic reviews))

### CINAHL

**Interface** - EBSCOhost Research Databases

**Search Screen** - Advanced Search

**Database** - CINAHL

((MH loneliness) OR (TX loneliness) OR ((TX lonely) OR (TX social isolation))) AND ((TX (digital* OR technol* OR sensor* OR robot* OR social media OR smartphone* OR smart phone* OR phone* OR online OR ipad* OR computer* OR elctronic* OR Web)) OR (MH technology) OR (MH robotics) OR (MH internet) OR (MH social media) OR (MH smartphone) OR (MH telephone) OR (MH computers, hand-held) OR (MH computers, portable) OR (MH world wide web))

### WEB OF SCIENCE

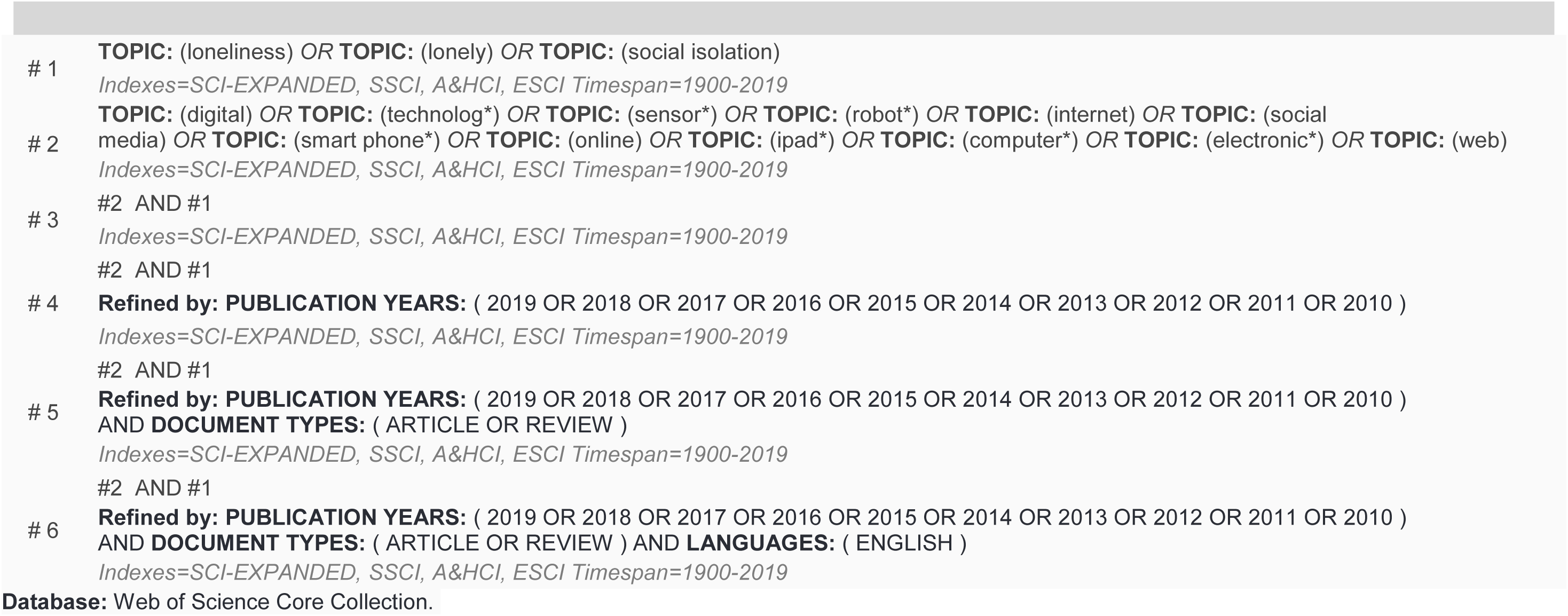

